# Applicability of Neighborhood and Building Scale Wastewater-Based Genomic Epidemiology to Track the SARS-CoV-2 Pandemic and other Pathogens

**DOI:** 10.1101/2021.02.18.21251939

**Authors:** Rachel R. Spurbeck, Angela T. Minard-Smith, Lindsay A. Catlin

**Affiliations:** Health Outcomes and Biotechnology Solutions, Battelle Memorial Institute, 505 King Ave, Columbus, OH 43201; National Security Bioscience Center, Battelle Memorial Institute, 505 King Ave, Columbus, OH 43201

**Keywords:** Wastewater based epidemiology, SARS-CoV-2, targeted genome sequencing, mutation analysis, RNAseq, viral load

## Abstract

The benefits of wastewater-based epidemiology (WBE) for tracking the viral load of SARS-CoV-2, the causative agent of COVID-19, have become apparent since the start of the pandemic. However, most sampling occurs at the wastewater treatment plant influent and therefore can only monitor SARS-CoV-2 concentration and spread within the entire catchment, which can encompass multiple municipalities. Furthermore, most WBE only quantifies the virus, and therefore miss crucial information that can be gained by sequencing SARS-CoV-2. Here we demonstrate feasibility of sampling at the neighborhood or building complex level using a mix of quantitative polymerase chain reaction (qPCR) and targeted sequencing to provide a more refined understanding of the local dynamics of SARS-CoV-2 strains. When coupled with the higher-level treatment plant samples, this creates an opportunity for health officials to monitor the spread of the virus at different spatial and temporal scales to inform policy decisions.

Here we demonstrate the feasibility of tracking SARS-CoV-2 at the neighborhood, hospital, and nursing home level with the ability to detect one COVID-19 positive out of 60 nursing home residents. The viral load obtained was correlative with the number of COVID-19 patients being treated in the hospital. Sequencing of the samples over time demonstrated that nonsynonymous mutations fluctuate in the viral population, and wastewater-based sequencing could be an efficient approach to monitor for vaccine or convalescent plasma escape mutants, as well as mutations that could reduce the efficacy of diagnostics. Furthermore, while SARS-CoV-2 was detected by untargeted RNA sequencing, qPCR and targeted whole genome amplicon sequencing were more reliable methods for tracking the pandemic. From our sequencing data, clades and shifts in mutation profiles within the community were traceable and could be used to determine if vaccine or diagnostics need to be adapted to ensure continued efficacy.

**Graphical Abstract:** 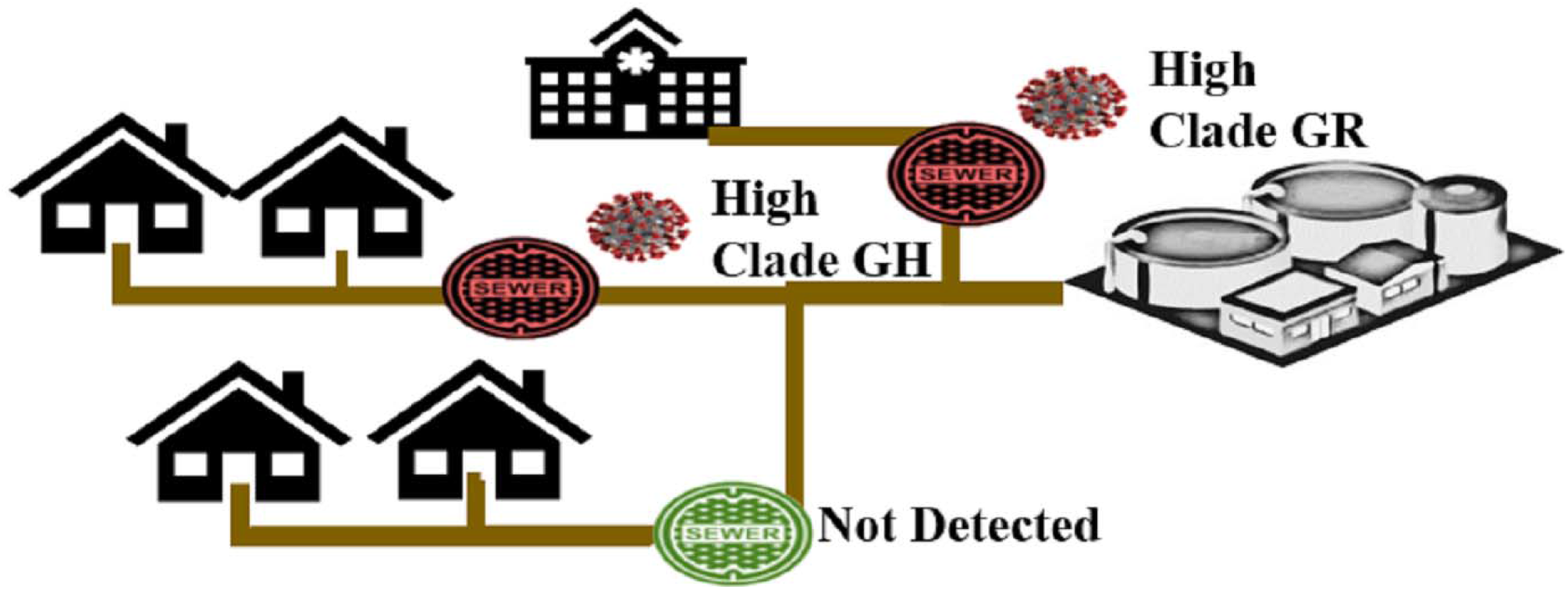

**Highlights:**

- Neighborhood or building level wastewater analysis accurately detects SARS-CoV-2
- SARS-CoV-2 was detected in wastewater from one infected person out of 60 residents
- Total RNAseq did not accurately detect SARS-CoV-2 in wastewater samples.
- Targeted whole genome sequencing of wastewater samples identified Spike mutations.

## 1.1 Introduction

Wastewater based epidemiology (WBE) is a rapidly growing field for the surveillance of pathogen load within human populations. Since it was discovered that SARS-CoV-2, the causative agent of COVID-19, can be quantified from feces (Wu et al., 2020b), WBE has been adapted to tracking the pandemic. Indeed, quantitative polymerase chain reaction (qPCR)-based wastewater testing has been implemented by several countries and states to track the COVID-19 pandemic by monitoring the viral load in wastewater (Achak et al., 2020; Ahmed et al., 2020; Bivins et al., 2020; Crits-Christoph et al., 2021; Foladori et al., 2020; Gonzalez et al., 2020; Gormley et al., 2020; Hamouda et al., 2020; Haramoto et al., 2020; Hart and Halden, 2020; Kitajima et al., 2020; Medema et al., 2020; Murakami et al., 2020; Weidhaas et al., 2020; Westhaus et al., 2021; Wu et al., 2020a). To maximize the information gathered from wastewater-based testing, we propose that targeted sequencing should be used in tandem with qPCR to determine the viral load and identify the dominant viral strain in circulation, including mutations that could affect diagnostic or therapeutic strategies. Furthermore, WBE implemented within catchments at the neighborhood or building level will enable a more refined map of COVID-19 hotspots.

WBE has gained popularity during the COVID-19 pandemic due to its ability to track the amount of the virus in a population using non-invasive sampling techniques (Murakami et al., 2020). Most studies sample influent to the wastewater treatment plant, which provides the ability to monitor the entire catchment, but does not have the resolution to gain a more detailed understanding of virus hotspots within that catchment. Hotspot information would be helpful for contact tracers to focus on neighborhoods, or for COVID-19 conscious individuals to determine what part of town is safest to go grocery shopping. The objectives of the current study were to demonstrate the application of WBE to track the spread of SARS-CoV-2 within a city and develop analytical pipelines to maximize information collected from these samples. In this work, wastewater was sampled from manholes located at mapped sewer junction points in the same city to provide a refined picture of COVID-19 prevalence in those communities.

WBE primarily focuses on viral load as the measure used to track SARS-CoV-2 prevalence and spread, but the technology exists for obtaining a more nuanced view of the viral strains and mutations that are in the samples. In this work, we explored not only qPCR to determine the viral load, but also targeted whole genome sequencing of SARS-CoV-2, and untargeted RNA sequencing to determine if SARS-CoV-2 can be detected without enrichment along with other pathogens of public health concern. Ideally, a wastewater-based tracking method should be optimized to understand all pathogen dynamics over time in a population. This can be achieved by non-targeted sequencing methods.

The sampling locations in this study were chosen specifically to enable comparison of viral load in wastewater to known case load or resident population numbers mapped to the sewer junction points. Two manholes used for sampling were located directly outside of major hospitals known to be treating COVID-19 patients, one was located outside of a nursing home, and a fourth manhole was located downstream of a residential neighborhood. The neighborhood also has a known number of residents, but an unknown number of COVID-19 cases. Therefore, the data collected could inform an estimated viral load in the community which may be correlated to the number of cases. Furthermore, two different RNA extraction methods and two sequencing approaches were used and evaluated to determine the most viable method to provide as much information as possible from each sample.

## 1.2 Materials and Methods

### 1.2.1 Wastewater sample collection

One-liter sample collections were conducted at four specific locations in Toledo, OH from manhole access once a week for three weeks. The first week of sampling, at three locations, the sample collections were grab samples due to low flow. The fourth location, Promedica Toledo Hospital, was collected as a 24-hour composite using an autosampler (ISCO 3700C). The subsequent collections at all locations were 24-hour composite samples. In low flow situations, a bucket was used to collect wastewater prior to use of the autosampler to ensure enough volume was present for the composite collection. All samples were collected between 10 AM and 12 PM EST and placed on ice in coolers during transportation to the analytical laboratory. As back up during analysis, 250 mL of each wastewater sample was stored. The following sample metadata was gathered each week during each sample collection: time of collection, method of collection, pH of samples, ambient temperature, and water temperature at time of collection.

### 1.2.2 Viral particle concentration

To encourage detachment of virions from organic material in the wastewater, 107 mL of glycine buffer (0.05M glycine, 3% beef extract, pH 9.6) was added to 750 mL of wastewater and mixed for one minute (Vlok et al., 2019). The samples were then centrifuged at 8,000 x g for 30 minutes. The supernatant was filtered through 0.22 micron filters to remove bacteria, fungi, or eukaryotic cells present in the samples. The virions were precipitated from the filtrate using a PEG-8000/NaCl solution ((PEG/NaCl) 320g/L PEG-8000, 70 g/L (w/v) NaCl) at a ratio of one-part PEG/NaCl to three parts filtrate (Wu et al., 2020a). Mixtures were gently stirred for 12 hours at 4°C. Precipitated virions were collected by centrifugation for 90 minutes at 13,000 x g at 4°C. Pellets were resuspended in 600 µL 1x PBS. The volume changes throughout processing were captured to enable back calculation of total viral load.

### 1.2.3 Viral RNA extraction

Two methods were used for RNA extraction. First, RNA was extracted from the collected viral preparations using TRIzol Reagent (ThermoFisher, Cat# 15596026) following the manufacturer’s instructions. RNA quantity and quality were measured by NanoDrop. Second, viral RNA was extracted by QIAamp Viral RNA Mini Kit (Qiagen, Cat# 52904) following the manufacturer’s protocol.

### 1.2.4 qPCR

Each RNA sample was analyzed by reverse transcription quantitative polymerase chain reaction (RT-qPCR) targeting the nucleocapsid gene segment (N1) to determine the SARS-CoV-2 viral load. Briefly, 5 µL of 4X TaqMan Fast Viral One-Step Master Mix (ThermoFisher, Cat# 4444432) 0.33 µL 60X custom TaqMan Gene Expression Assay primer/probe mix (Forward primer 5’ -GAC CCC AAA ATC AGC GAA AT-3’ ; Reverse primer 5’ -TCT GGT TAC TGC CAG TTG AAT CTG-3’ ; Probe 5’ -FAM-ACC CCG CAT TAC GTT TGG TGG ACC-NFQ-MGB-3’) (ThermoFisher, Cat# 4331348), 9.67 µL molecular biology grade water, and 5 µL sample were combined for a 20 µL reaction volume. Each plate contained an 8 standard (SARS-CoV-2 synthetic RNA from Biosynthesis, Inc.) serial dilution (1E7 to 1E0) run in triplicate, no template control in triplicate, and samples were in duplicate. Plates were run according to the following thermal cycling parameters: 50°C for 5 min, 95°C for 20 sec, and 40 cycles of 95°C for 3 sec followed by 60°C for 30 sec in an ABI 7500 fast quantitative thermocycler.

### 1.2.5 RNA Virus metagenomics

Total RNA was sequenced from each sample. RNA libraries were prepared using the Rapid RNAseq kit (Swift, Cat# R2096) from Swift Biosciences and sequenced on an Illumina NextSeq using two NextSeq 500/550 High Output Kits v2.5 (150 Cycles, 75x 75 bp). No template control libraries were prepared alongside RNAseq samples, but not sequenced. Data were analyzed following the pipeline outlined in the flowchart depicted in Figure 1.

**Figure 1.**
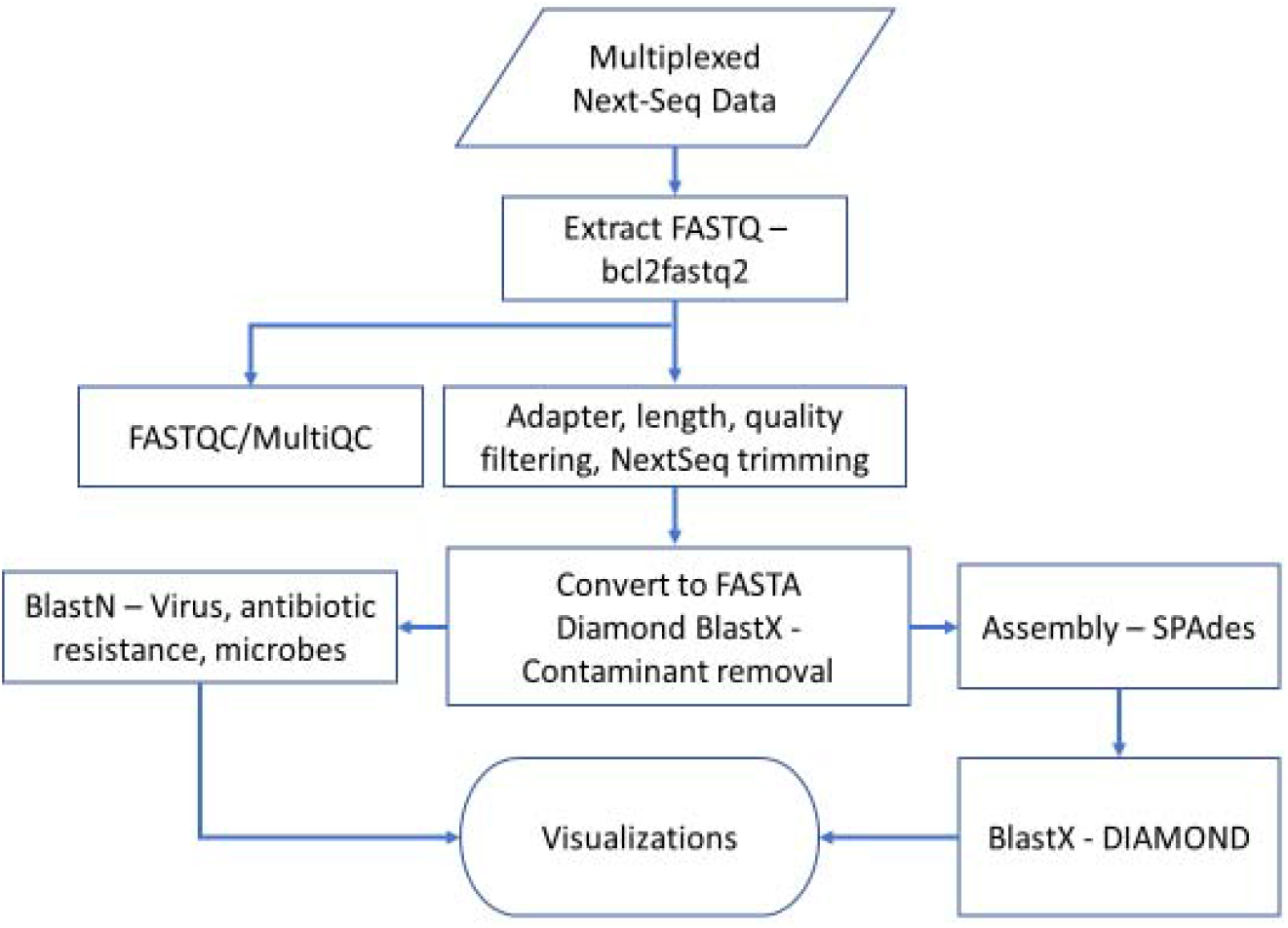
Processing flowchart for Total RNA-Seq data. RNA-Seq data was obtained from runs on an Illumina NextSeq instrument. Files will demultiplexed, quality checked, and trimmed. Sequences were assembled using SPAdes into contigs and then compared to a known reference sequences in the refseq database using BlastN or BlastX-DIAMOND to identify pathogens in the wastewater samples.

### 1.2.6 COVID-19 amplicon sequencing

Samples were tested specifically for SARS-CoV-2 by amplicon sequencing using the SNAP SARS-CoV-2 Kit (Swift, Cat# SN-5×296 and COVG1-96) from Swift Biosciences. Briefly, first strand cDNA was synthesized from 11 µL of total RNA from each sample using the Superscript IV First Strand Synthesis System with random hexamers (Invitrogen, Cat# 18091050) following the manufacturer’s instructions. The resulting cDNA (10µL) was used as template for the SARS-CoV-2 Kit following the manufacturer’s instructions. Prior to sequencing, the samples were quantified by Qubit HS DNA kit (Invitrogen, Cat# Q32851), pooled, and sequenced on an Illumina MiSeq 300 cycle V3 flow cell (Illumina, Cat# MS-102-2002). Samples were batched by week to enable high depth of coverage for single nucleotide polymorphism analysis. The amplicon data were aligned to the NCBI Reference Sequence NC_045512.2 (Severe acute respiratory syndrome coronavirus 2 isolate Wuhan-Hu-1, complete genome) and analyzed using the pipeline provided by Swift Biosciences. A positive control and a no template control were processed and quantified. The no template control was not sequenced, but the positive control was sequenced demonstrating full genome coverage of reference sequence NC_045512.2.

## 1.4 Results

### 1.4.1 Sampling site selection

Unlike most wastewater epidemiology studies which sample at the influent going into the wastewater treatment plants servicing a particular region of a city, our project studied the biological components in raw sewage derived from manholes in four specific locations in the city of Toledo, Ohio. The sites were carefully chosen through discussion with the city wastewater management team and the Toledo-Lucas County Public Health Department to ensure the sites were in locations with a high probability of SARS-CoV-2 circulation. Two sites were the direct effluent from hospitals known to be actively treating COVID-19 patients at the time of the study, Promedica Toledo Hospital (P, Figure 2A), and Mercy Health Heart and Vascular Institute at St. Vincent Medical Center (V, Figure 2B). A third site was outside a nursing home, Continuing Healthcare Solutions (N, Figure 2C) and a fourth was outside a residential neighborhood, Cresthaven (Ragan Woods, in the western portion of the Southwyck neighborhood, C, Figure 2D).

**Figure 2.**
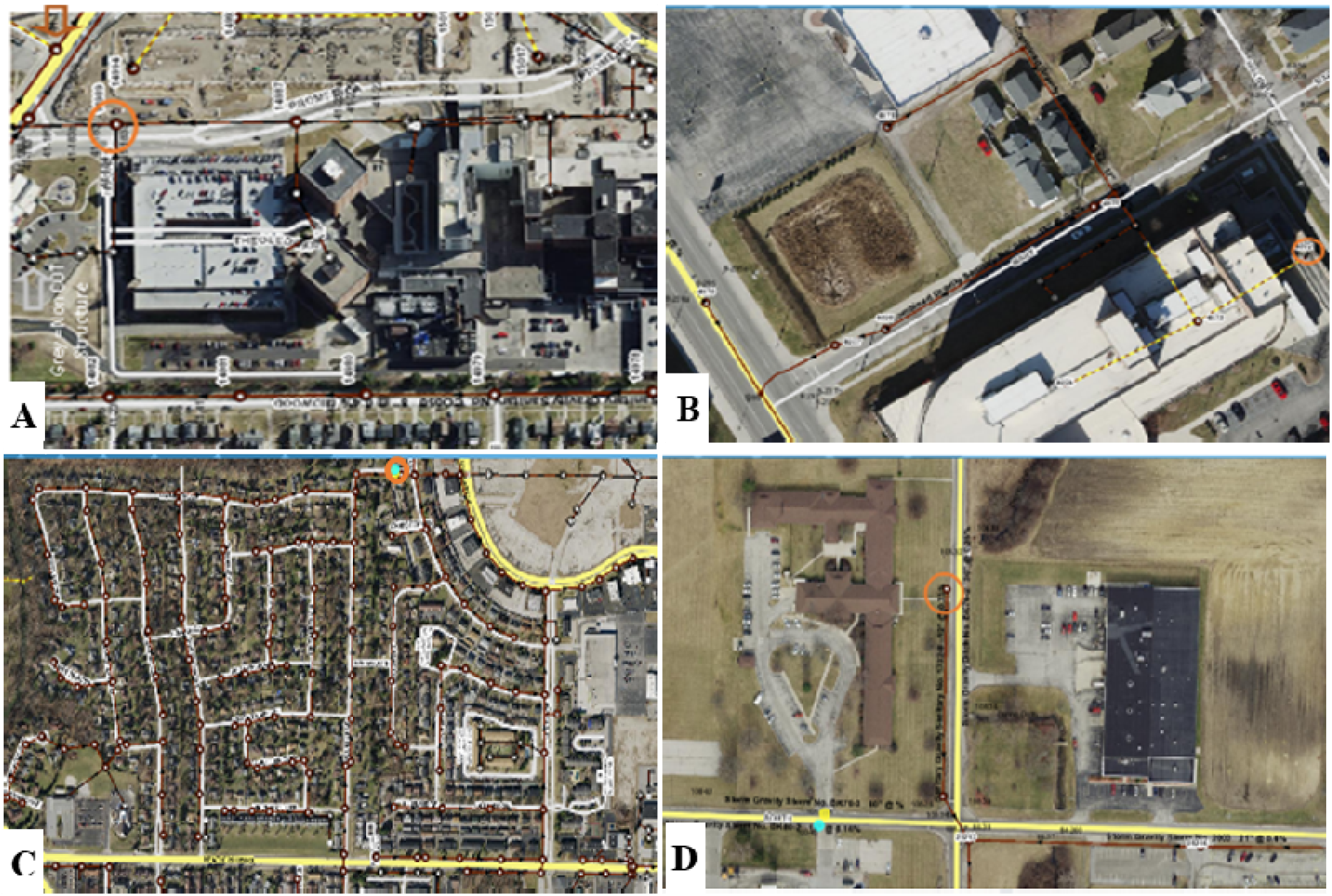
Sampling sites within Toledo, Ohio. Orange circles indicate manholes sampled. Brown lines indicate sewer lines. A. Promedica Toledo Hospital. B. St. Vincent’s Heart Center. C. Cresthaven Neighborhood. D. Continuing Healthcare of Toledo Nursing home.

### 1.4.2 Known case load at sampling sites by week and qPCR detection of SARS-CoV-2

The RNA extracted by the TRIzol reagent did not show evidence of any SARS-CoV-2 in the samples, however, the QIAamp Viral/Pathogen RNA preparations produced detectable levels of SARS-CoV-2 by qPCR in 9 of the 12 samples (Table 1). For comparison, the percentage of known COVID-19 cases in the hospitals and nursing home were determined by dividing the number of known COVID-19 positive patients by the total number of inpatients, and a weak positive correlation is observed between the number of infected and viral load in the wastewater.

**Table 1.** Percent of positive cases compared to the viral load detected in wastewater.

|  | 7/14/20 |  | 7/22/20 |  | 7/28/20 |  |
| --- | --- | --- | --- | --- | --- | --- |
| *Site* | *Percent Positive (%)* | *Viral Load (gc/mL)* | *Percent Positive (%)* | *Viral Load (gc/mL)* | *Percent Positive (%)* | *Viral Load (gc/mL)* |
| *C* | *Unknown* | 39.1 | *Unknown* | 4.5 | *Unknown* | 2.9 |
| *N* | 0 | ND | 0 | ND | 2 | 0.6 |
| *P* | DNP | 2.3 | DNP | 1.0 | DNP | ND |
| *V* | 8 | 17.4 | 12 | 9.0 | 15 | 110.6 |

DNP = Data not provided, ND = not detected

### 1.4.4 Non-targeted RNAseq detection of SARS-CoV-2

In theory, RNAseq should afford direct SARS-CoV-2 signature detection in wastewater without requiring enrichment. However, despite following viral concentration protocols, a large amount of the sequence data was found to map to other organisms such as bacteria, bacteriophage, and eukaryotic organisms (Figure 3). Reads mapping to SARS-CoV-2 were detected in only five out of the twelve samples, with at most 3 reads mapping to the viral genome, despite there being an average of 11M raw reads per sample and 3M filtered reads per sample. SARS-CoV-2 was detected in Cresthaven samples collected week 1 (3 reads) and week 2 (3 reads), and St. Vincent’s samples collected in week 1 (1 read), week 2 (2 reads), and week 3 (2 reads). This low coverage is not sufficient for confidence in reporting presence of the pathogen at the local level. Due to the relatively small genome and other larger genomes present in the wastewater, the SARS-CoV-2 reads were likely obscured. We therefore recommend tracking the pandemic using a local surveillance level (residential neighborhoods or buildings) combined with a targeted enrichment method, such as amplicon-based sequencing.

**Figure 3.**
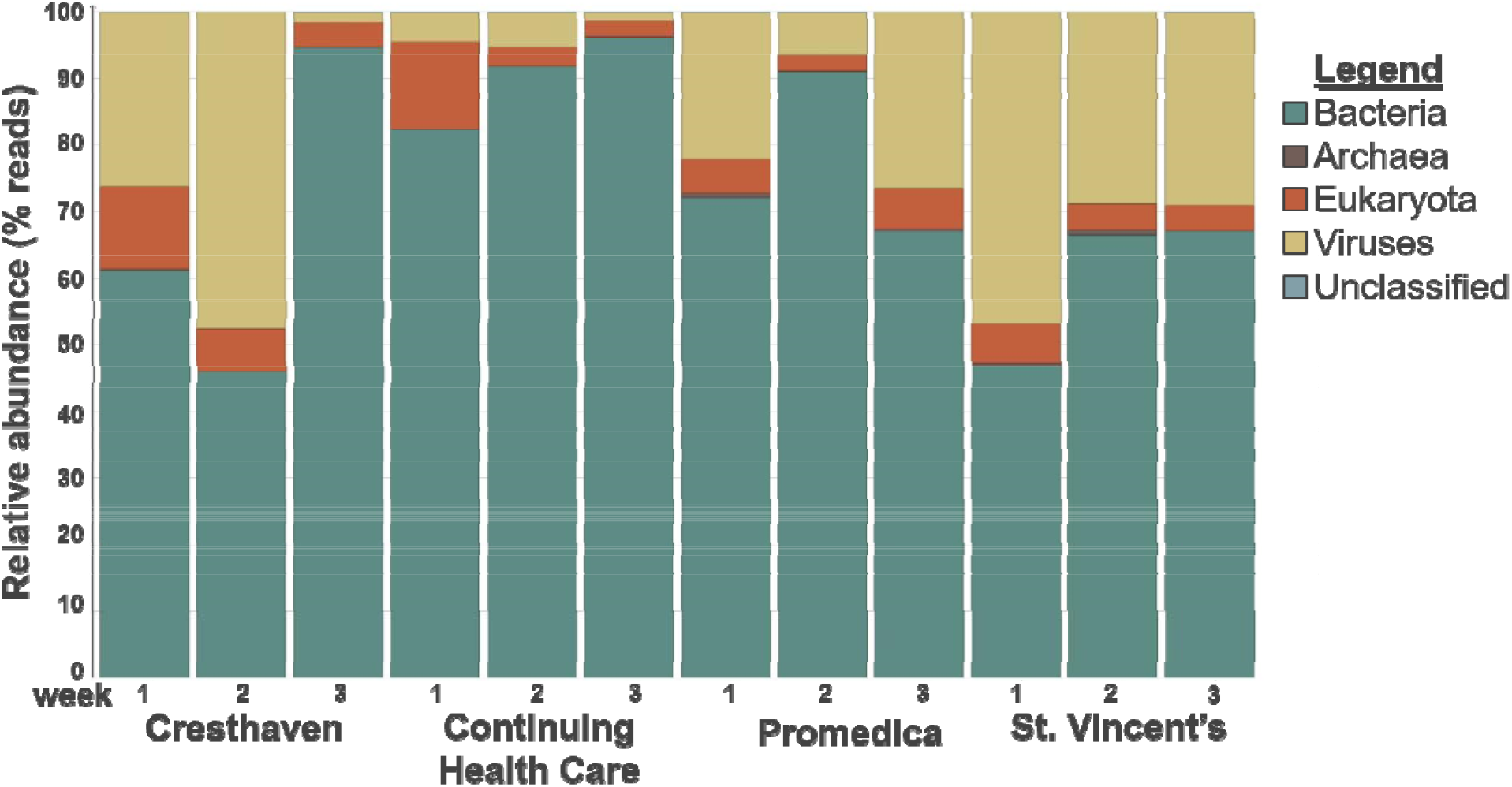
Relative abundance of viruses, bacteria, archaea, and eukaryotes identified by total RNA sequencing of wastewater. Collection dates are indicated as 1 (7/14/20), 2 (7/21/20), or 3 (7/28/20).

Despite removal of microorganisms and enrichment for viral particles, a large portion of the sequencing data (51-90%) mapped to bacteria, and a fraction of the data (1-19%) mapped to macroorganisms, which demonstrates our viral preparation also included RNA from lysed cells (Figure 3). Although not the target of this study, the sequenced microbial RNA identified several commensal species that are nonpathogenic to their hosts, as well as infectious and opportunistic pathogens (Figure 4). While not pertaining to the COVID-19 pandemic, this finding demonstrates that other pathogenic organisms can be identified and tracked using wastewater sequencing, and therefore non-targeted wastewater sequencing could be continued after the pandemic wanes to improve our understanding of pathogen distribution within local communities and healthcare facilities. Several pathogenic categories of interest were detected in the hospitals, nursing home, and community, including microbes associated with nosocomial infections, gastroenteritis, cystitis, pneumonia, septicemia, and tetanus (Figure 5) from assembled contigs. The etiological agents identified for nosocomial infections included the *Acinetobacter* species: *A. baumannii A. junii SH205*, and *A. Iwoffii SH145. Campylobacter jejuni subsp. jejuni BH-01-0142*, a known cause of gastroenteritis, was identified in the Cresthaven neighborhood, the Continuing Healthcare nursing home, and St. Vincent’s Hospital. The causative agent of cystitis identified in the first week of testing at Continuing Healthcare was *Escherichia coli* UTI89. This data suggests that WBE can be utilized to track contagions outside of a pandemic and could be a beneficial tool for future epidemiological and biosurveillance studies.

**Figure 4.**
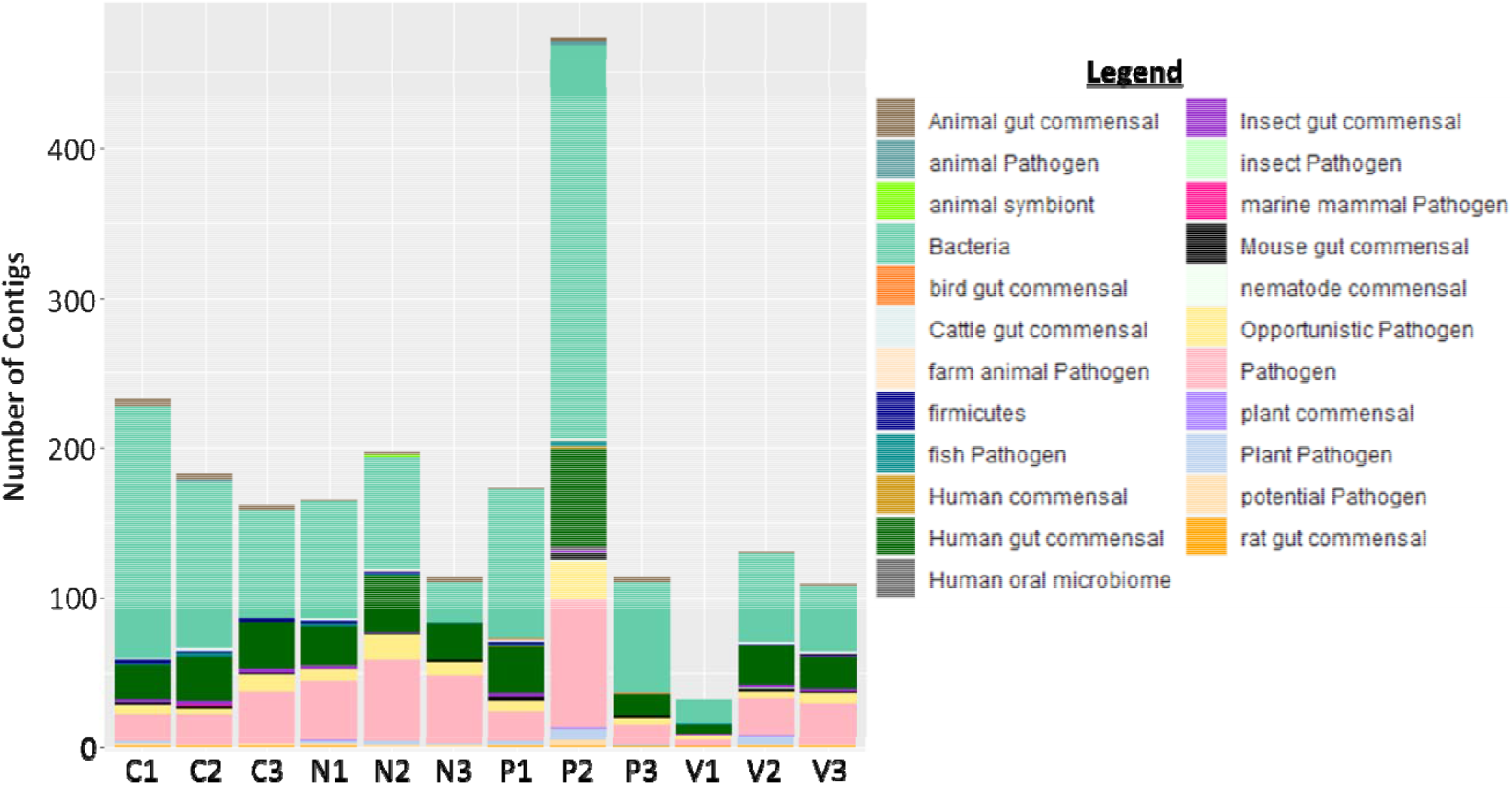
Ecological niches associated with the microorganisms identified by total RNA sequencing identified several human, animal and pl ant pathogens, as well as potential and opportunistic pathogens. Sample sites are indicated as Nursing home (N), St. Vincent’s Heart Center (V), Promedica Toledo Hospital (P), and the Cresthaven Community (C). Collection dates are indicated as 1 (7/14/20), 2 (7/21/20), or 3 (7/28/20).

**Figure 5.**
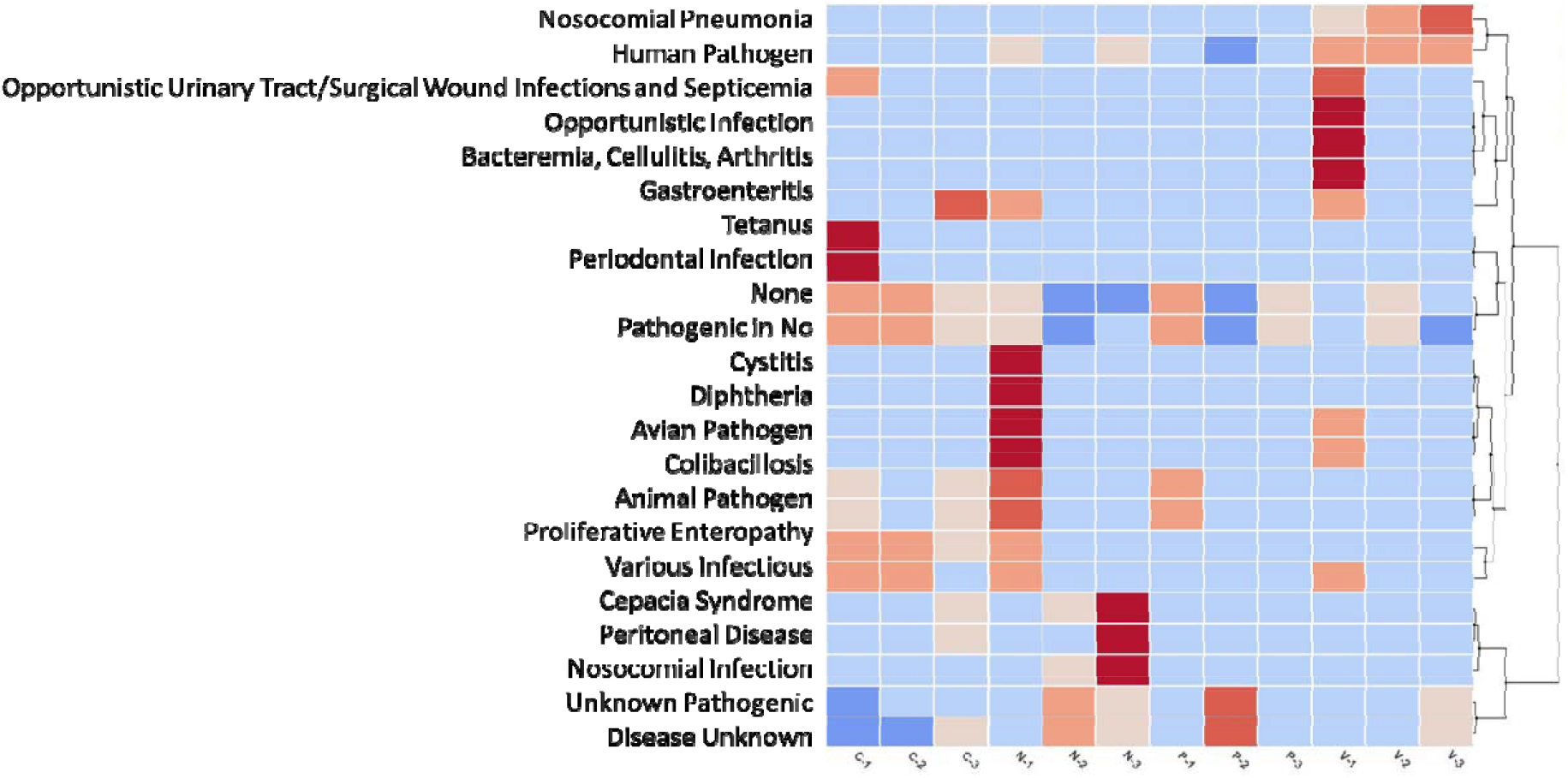
The abundance of pathogens identified by total RNA sequencing varied by sampling site and over time. Notably, pathogens associated with nosocomial infections were found in the effluent from the nursing home (N) and St. Vincent’s Heart Center (V), but not in Promedica Toledo Hospital (P) or the Cresthaven Community (C). weeks of collection are indicated as 1 (7/14/20), 2 (7/21/20), or 3 (7/28/20).

### 1.4.5 Amplicon sequencing

The whole genome of SARS-CoV-2 was enriched from the wastewater samples by targeted amplicon sequencing, enabling single nucleotide polymorphisms (SNP) and insertion/deletion mutations (indels) to be identified when aligned to the reference strain SARS-CoV-2 Genome Wuhan HU-1. Table 2 presents non-synonymous mutations identified in the Toledo samples that are unique (previously unobserved) or existing mutations (previously observed). Of the twelve samples analyzed in this study, nine were COVID-19 positive based on qPCR. Sequencing the positive samples, only five produced full genomes with enough coverage for confident calls: weeks 1 and 2 from the Cresthaven neighborhood and weeks 1, 2, and 3 from St. Vincent’s hospital (Table 2 and Figure 6). The mutations identified enabled Clade classification. Community samples taken from the Cresthaven neighborhood fell into Clade GH, which carries the Spike D614G mutation. St. Vincent’s data showed several mutations at variable penetrance indicating multiple strains are present in these samples (Figure 6), therefore the Clade was described as Other. Despite having multiple strains present, all three weeks had SARS-CoV-2 sequences carrying the Spike D614G mutation.

**Table 2.** Consensus sequences identified nonsynonymous mutations and categorized the strains by clade.

| Sample | Genome coverage (%) | Length (AA) | #Mutations (%) | Unique Mutations | Existing Mutations | Clade |
| --- | --- | --- | --- | --- | --- | --- |
| **C-1** | 97.43% | 9688 | 13 (0.13%) | NSP14_V115I | NSP2_T85I, **NSP3_M1788I**, NSP5_L89F, NSP12_P323L, NSP14_N129D, NSP16_R216C, Spike_D614G, NS3_Q57H, NS3_G172V, NS8_S24L, N_P67S, N_P199L, | GH |
| **C-2** | 86.09% | 8262 | 13 (0.16%) | NSP13_G170R<br>NSP14_S418C<br>Spike_N370K | NSP2_T85I, NSP5_L89F, NSP14_N129D, NSP16_R216C, Spike_D614G, NS3_Q57H, NS3_G172V, NS8_S24L, N_P67S, N_P199L | GH |
| **C-3** | 1.88% | 61 | -- | -- | -- | -- |
| **N-3** | 8.87% | 752 | 1 (0.13%) |  |  | -- |
| **P-1** | 30.66% | 2459 | 4 (0.16%) |  |  | GR |
| **P-2** | 26.31% | 2164 | 7 (0.32%) |  |  | GH |
| **V-1** | 92.78% | 9642 | 14 (0.15%) | NSP3_A416V<br>NSP6_K151R | NSP2_T85I, NSP3_M1788I, NSP5_L89F, NSP12_P323L, NSP14_N129D, NSP16_R216C, Spike_D614G, NS3_Q57H, **NS3_G172V**, NS8_S24L, N_P67S, **N_P199L** | Other |
| **V-2** | 91.66% | 8826 | 15 (0.17%) | NSP3_A1262S<br>NSP3_N1263T<br>NSP14_S418C<br>M_V10I | **NSP1_H110Y**, NSP2_T85I, NSP3_M1788I, NSP5_L89F, NSP12_P323L, NSP14_N129D, NSP16_R216C, Spike_D614G, NS3_Q57H, NS8_S24L, N_P67S | Other |
| **V-3** | 99.57% | 9710 | 9 (0.09%) | NSP3_F1107L<br>NSP14_N129B | NSP2_T85I, NSP3_M1788I, NSP12_P323L, NSP16_R216C, Spike_D614G, **NS3_G172V**, **M_A2V** | Other |

\*Gray rows indicate samples with partial genome coverage, which may not reliably call the clade. Bold mutations are ones that were not present in all weeks at the same site.

A week-to-week comparison of Cresthaven samples yielded highly similar existing nonsynonymous mutation loads with ten mutations observed in both weeks. In addition, different unique nonsynonymous mutations were observed, and one existing nonsynonymous mutation was lost from week 1 to week 2. All three weeks from St. Vincent’s had five existing nonsynonymous mutations in common (NSP2_T85I, NSP3_M1788I, NSP12_P323L, NSP16_R216C, and Spike_D614G). However, St. Vincent’s weeks 1 and 2 had an additional five existing nonsynonymous mutations in common (NSP5_L89F, NSP14_N129D, NS3_Q57H, NS8_S24L, and N_P67S), demonstrating that nonsynonymous mutation load has changed over time.

Similarly, the number of SNPs and indels in the whole genome sequencing data changed over time at each site (Table 3). The three genes most targeted by diagnostics were prone to both SNPs and indels, demonstrating the importance of continuous monitoring by sequencing as the pandemic continues. Over the three weeks of this study, the viral population present in the effluent from St. Vincent’s underwent the most striking changes, with the total number of SNPs and indels increasing each week. Importantly, two deletions found in the S protein in the third week of sampling (7/28/20) at St. Vincent’s was represented in 34% of the data; a deletion of a full codon, ΔACC23493-23496, found in 9% of the data, and one was found at 25%: ΔG24696.

**Table 3.** SNP frequency at genes targeted for diagnostics or therapeutics and number of indels.

|  | *Cresthaven* |  | *St. Vincent's* |  |  |
| --- | --- | --- | --- | --- | --- |
|  | Week 1 | Week 2 | Week 1 | Week 2 | Week 3 |
| *orf1ab* | 0.056% | 0.038% | 0.042% | 0.103% | 0.211% |
| *S* | 0.078% | 0.078% | 0.026% | 0.052% | 0.235% |
| *N* | 0.159% | 0.159% | 0.317% | 0.635% | 0.238% |
| *all other* | 0.170% | 0.085% | 0.142% | 0.227% | 0.340% |
| *#indels* | 0 | 2 | 3 | 9 | 17 |

## 1.5 Discussion

The objective of this work was to first demonstrate the utility of wastewater surveillance at the neighborhood or hospital level and then to identify the best method for tracking SARS-CoV-2 and other pathogens to enable a population-based approach to pandemic biosurveillance. Our data have demonstrated that while total RNAseq, targeted whole genome sequencing, and qPCR methodologies can be used to detect SARS-CoV-2 in wastewater, a combination of qPCR to identify viral load and targeted whole genome sequencing to detect mutations provides deeper insights to support public health decisions during pandemic surveillance than qPCR alone. RNAseq, while identifying other pathogens present in the samples such as microbes that cause nosocomial infections, periodontitis, gastroenteritis, pneumonia, or cystitis, did not recover enough reads mapping to SARS-CoV-2 to detect the viral pathogen with any confidence. However, the non-targeted surveillance technique would effectively alert hospitals and long-term care facilities to potential issues with nosocomial pathogens or residents that may have infections that need to be treated and can otherwise go undiagnosed such as cystitis due to asymptomatic or recurrent infections. Use in community settings would alert neighborhoods to potential norovirus or *Salmonella* outbreaks and help public health officials pinpoint restaurants or grocery stores selling contaminated products.

Utilizing amplicon sequencing to target and sequence the full SARS-CoV-2 genome enabled mutation analysis of the viral population identified in the wastewater samples. As demonstrated recently, new strains of SARS-CoV-2 are emerging which are hyper transmissible, and potentially could carry mutations in the Spike protein that could be vaccine or convalescent plasma therapy escape mutants (England, 2020; Kemp et al., 2020; Tu et al., 2021). Also, the Spike, N, or Orf1ab genes are diagnostic targets so any SNP or indel identified in these genes that could affect primer or probe binding of a diagnostic test are of high interest and need to be reported to the scientific community to enable improved diagnostics. Indeed, several mutations have been identified that affect different COVID-19 diagnostic tests (Wang et al., 2020). Furthermore, nonsynonymous mutations in the Spike protein could indicate the potential for vaccine escape and will need to be characterized to understand impact on vaccine efficacy. Sequencing from wastewater samples within a city or at hospital effluent can aid in surveillance of these mutations and reduces the burden on supplies, as instead of sampling, quantifying and sequencing SARS-CoV-2 in individuals, wastewater enables population sampling, reducing the number of assays and reagents used during surveillance.

With both convalescent plasma and the vaccines targeting the spike protein, it is imperative that nonsynonymous mutations be monitored in this gene. Indeed, it has been demonstrated that spike protein mutations increase in SARS-CoV-2 as a patient is treated with convalescent plasma, indicating the antibodies are placing evolutionary pressure on the virus that selects for mutations in this therapeutic target (Kemp et al., 2020). All Toledo wastewater samples sequenced carried the Spike G614D mutation, which may confer higher transmissibility to the virus, and is characteristic of clade G. Indeed, three of the sequenced samples were categorized as GH and one as GR. In a study in Asia, clades GH and GR were found to contain high levels of SNPs, indicating a lot of diversity in these strains (Sengupta et al., 2020). Mutation analysis of wastewater samples collected in Toledo OH during July 2020 did not find signature for the two SARS□CoV□2 clade 20C/G variants that were later identified in Columbus, OH (Tu et al., 2021). Likewise, the strain UK□B.1.1.7 (clade 20I/501Y.V1)(England, 2020; Galloway et al., 2021), also thought to be hyper transmissible and bearing mutations in the S protein, was not present in July 2020 Toledo, OH wastewater samples. However, another spike mutation, Spike_N370K, was observed in week 1 of the Cresthaven neighborhood, and two deletions (ΔACC23493-23496 and ΔG24696) were observed in week 3 at St. Vincent’s. Therefore, wastewater-based sequencing of SARS-CoV-2 can identify spike protein mutations which may impact transmission and response to the vaccine or convalescent plasma therapy.

Most wastewater-based epidemiology takes place at the influent to a wastewater treatment facility, providing a large catchment that encompasses multiple municipalities. In contrast, our work focused on sampling effluent from a neighborhood, two hospitals, and a nursing home. While these sites provided smaller population sizes and can have flow rate issues (some of our sample locations could not provide 24-hour composites due to low flow), sampling at regions within a city provides more refined information on where strains are circulating in the community. For example, strains circulating in Cresthaven were all in the clade GH, while a strain identified in Promedica Toledo Hospital belonged to clade GR, and strains at St. Vincent’s were categorized as Other. While all strains bore some mutations in common, each week’s sample had mutations unique to that sampling, demonstrating that mutations are common in SARS-CoV-2, and several strains are present in one city.

While population size is a concern, qPCR from effluent of our smallest population, that of the nursing home Continuing Healthcare, was able to detect the single case of COVID-19 in week 3, from a population of 60 residents. Since Ohio has implemented weekly testing of nursing home residents, this detection was confirmed by the public health department’s data. The case load data from St. Vincent’s hospital demonstrated an increase in hospitalized COVID-19 cases over time, and our qPCR data correlates with this trend, indicating that with more sampling, a predictive model could be developed and validated to identify an approximate number of infected people present in the catchment over time. Using data from the ongoing study, we are currently working to develop and validate candidate models.

Another highly discussed aspect of wastewater-based epidemiology is the degradation of target viral RNA during its transit through the sewer system. While SARS-CoV-2 RNA is intact enough at the inlet to the wastewater treatment plant for qPCR identification, sequencing samples taken closer to the source, such as the effluent from a building or a neighborhood may provide less degraded samples for whole genome sequencing, and therefore provide a better representation of the viral mutations in circulation. In the future, we would like to compare RNA quality of samples from different locations within a catchment to optimize viral sequencing.

### 1.5.1 Conclusions

This study has demonstrated that wastewater epidemiology at the local level can be used to determine the viral load in a neighborhood or building and identify strain variants in circulation. Total RNAseq from wastewater samples, while effective in identifying bacterial pathogens in circulation, was not able to consistently detect SARS-CoV-2 for biosurveillance purposes, due to the low abundance of the viral RNA in comparison with all other RNA in the sample. Therefore, it is recommended that a combination of qPCR and targeted sequencing of SARS-CoV-2 be utilized to track the pandemic and provide early warning of new strains circulating in the population which could otherwise evade detection by diagnostics or indicate vaccine escape mutants. The use of wastewater in to track mutations is limited by the methods to bioinformatically deconvolute the viral sequences, and therefore in this work we presented the mutations identified without separating the sequences into individual genomes. Future directions will incorporate long read sequencing or linked read sequencing to enable deconvolution into individual viral strains. Furthermore, a comparison of genomic sequences from samples at wastewater treatment plants versus the local manholes utilized in this study would determine if the time between viral shedding in waste and collection impacts viral RNA integrity limiting the use of genomic sequencing to building or neighborhood wastewater samples for high quality full genome sequences.

## Data Availability

data utilized in the study can be made available upon request.

## Acknowledgements

The authors would like to thank Dennis McIntyre, Great Lakes Environmental Center for sample collection; Angela Tucker, City of Toledo, Division of Environmental Services for help with sample site identification and access to sewers; and Dr. Jennifer Gottschalk, Toledo-Lucas County Public Health Department, and St. Vincent’s Heart Center for providing COVID-19 case data. We would also like to thank Drs. Jared Schuetter and Trevor Petrel for assistance with critique and editing the manuscript.

